# Molecular network analyses implicate death-associated protein kinase 3 (DAPK3) as a key factor in colitis-associated dysplasia progression

**DOI:** 10.1101/2021.09.21.21263916

**Authors:** Huey-Miin Chen, Justin A. MacDonald

## Abstract

Ulcerative colitis (UC) is a progressive disorder that elevates the risk of cancer development through a colitis-dysplasia-carcinoma sequence. Differential gene expression (DEGs) profiles of three UC clinical subtypes and healthy controls were developed for the GSE47908 microarray dataset [n = 15 (healthy controls), n = 20 (left-sided colitis), n = 19 (pancolitis), and n = 6 (colitis-associated dysplasia, CAD)] using limma R. Gene ontology (GO) enrichment analysis of DEGs revealed a shift in transcriptome landscape as UC progressed from left-sided colitis to pancolitis to CAD, from being immune-centric to being cytoskeleton-dependent. Hippo signaling (via Yes-associated protein, YAP) and Ephrin receptor signaling were the top canonical pathways progressively altered in concert with the pathogenic progression of UC. Molecular interaction network analysis of DEGs in left-sided colitis, pancolitis, and CAD revealed one pairwise line or *edge* that was topologically important to the network structure. This edge was found to be highly enriched in actin-based processes, and death-associated protein kinase 3 (DAPK3) was a critical member and sole protein kinase associated with this edge. DAPK3 is a regulator of actin-cytoskeleton reorganization that controls proliferation and apoptosis. Differential correlation analyses revealed a negative correlation for *DAPK3-YAP* in healthy controls which flipped to positive in left-sided colitis. With UC progression to CAD, the *DAPK3-YAP* correlation grew progressively more positive. In summary, DAPK3 was identified as a candidate gene involved in UC progression to dysplasia.

## 1. INTRODUCTION

Ulcerative colitis (UC) is a chronic, inflammatory bowel disease (IBD) that is confined to the mucosal layer of the large bowel, most commonly the rectum, and may extend proximally in a continuous fashion (1). This is a heterogenous and progressive disorder that has seen a substantial increase in global prevalence. The current framework of UC pathogenesis comprises environmental, genetic, immune, and microbiome factors that culminate at the perturbation of the mucosal barrier and prolonged mucosal inflammatory response (2). Consequently, UC is a clinically-, molecularly-, and genetically-heterogeneous disease that fosters varied disease course and mixed response to therapy (3). The genetic heterogeneity is highlighted in the analysis of 75,000 IBD cases and controls by Jostins and colleagues that identified 23 UC-specific risk loci with primary involvement in the regulation of epithelial barrier function and immune pathways (4). Genetic studies such as this promote the concept that UC disease susceptibility is a compilation of small effects/gene-alterations that is not shared by all patients.

UC patients are at greater risk of developing colitis-associated colorectal cancer (CAC) through an acceleration of the colitis-dysplasia-carcinoma sequence of cellular transformation (5). Mutational signature analysis of tissue and blood samples from CAC patients showed that the evolutionary trajectory of disease is initiated early in the colitis-dysplasia-carcinoma sequence (6). Furthermore, the extent of colitis has been recognized as an independent and most significant risk factor for CAC (7). In support of this, Bjerrum and colleagues provided transcriptional profiles of colonic mucosa from patients with varying extent of UC (8). A gene expression dataset (GSE47908) was generated from mucosal biopsies sampled from the left colon of patients with left-sided colitis, pancolitis, or colitis-associated dysplasia (CAD) plus healthy controls. The authors identified differential transcriptional profiles with primary component analysis (PCA) aligned with UC extent (i.e., areas of involvement) that were not inferred by potential covariates in the clinical data (i.e., age, years with disease, Mayo score, and medication). Their findings suggest that gene expression profiles of colitis-associated lesions obtained from patients with varied extent of UC can be mined to support the development of molecular panels that identify patients at high risk of developing dysplasia or CAC. However, additional network analyses and detailed interrogation of specific molecular participants were not provided in the Bjerrum study.

Elucidation of the mechanisms of colon carcinogenesis requires further investigation, and insight into the molecular events underpinning the progression of UC to colitis-associated colon cancer may be gained from the study of non-dysplastic colonic mucosa (6,8). The transcriptional dataset generated by Bjerrum and colleagues (8) provides an excellent resource to perform such an analysis. In this study, we performed differential expression analysis, pathway and network analyses, as well as differential correlation analysis on the GSE47908 dataset to determine how the extent of colitis impacts upon biological functions and regulatory pathways and to identify the key molecular factors that bridge the colitis-dysplasia progression. The results provide insight into the molecular events associated with colitis-dysplasia progression, which could be exploited for the development of biomarkers in non-dysplastic mucosa that identify the risk of dysplasia for UC patients.

## 2. METHODS

### 2.1. Data Processing

Data processing was completed using the R (v4.0.2) programming language, and all codes used in this study align with recommendations made by authors of R packages in their respective user’s guide, which can be accessed at https://bioconductor.org.

### 2.2. Differential Gene Expression Analysis

Log transformed microarray expression data for GSE47908 and microarray platform data for GPL570 (HG-U133_Plus_2; Affymetrix Human Genome U133 Plus 2.0 Array) were retrieved from the GEO database available at https://www.ncbi.nlm.nih.gov/geo/ with the R package GEOquery (9). The limma workflow (10) was used to detect differentially expressed transcripts between the UC clinical subtypes [left-sided colitis (n=20), pancolitis (n=19), colitis-associated dysplasia, CAD (n=6)], and healthy control (HC) samples (n=15). Specifically, the function |lmFit| was used to generate a linear model fit to the data matrix containing log expression values for GSE47908. Next, function |contrasts.fit| was used to compute estimated coefficients and standard error for a given set of contrasts (e.g., pancolitis vs. HC). Finally, function |eBayes| was used to compute log-odds of differential expression by empirical Bayes moderation of the standard errors towards a global value. All functions were operated with default settings.

Log-fold changes calculated by function |eBayes| for 54,675 transcripts, along with their false discovery ratio (FDR) and p-values, were uploaded to the Ingenuity Pathway Analysis (IPA) software (11). Some Affymetrix transcript identifiers remain unmapped by IPA; these genes were eliminated from the study, leaving 45,480 transcripts to be mapped. Due to redundancy of the GeneChip HG-U133 Plus 2.0 Array, this transcript pool included duplicate genes. To resolve duplicates, IPA Core Analyses were performed on the mapped transcripts, and transcript identifiers were consolidated using their log_2_FC measurement, by which, representative transcript was selected based on maximum absolute log_2_FC. This returned 21,475 ‘analysis-ready’ genes. To distinguish the differentially expressed genes (DEGs), the combined application of a stringent FDR threshold (q < 0.001) and a moderate fold change threshold |log_2_FC| > 0.75) was used. This reduced the number of false positives while maintaining the ‘ideal’ dataset size (200-3,000 genes) for subsequent IPA Core Analysis of gene expression data (11). Differential expression was then visualized via the EnhancedVolcano R package (12).

### 2.3. Functional Analysis of DEGs in UC disease subtypes

To compare DEGs and illustrate possible relationships between left-sided colitis, pancolitis, and CAD, a Venn diagram was first used to visualize the overlap of DEGs found in the three UC clinical subtypes. The lists of overlapping DEGs derived from the comparison of UC subtypes were then used as input data for gene ontology (GO) enrichment analysis, performed with the topGO R package (13). topGO was chosen owing to its *‘elim’* method that takes GO hierarchy into consideration when calculating enrichment. For the enrichment analysis, the background consisted of all genes assessed by the microarray platform GPL570, and annotation was completed with the R package org.Hs.eg.db (14); GO terms with <10 annotations were excluded for interpretability. The degree of enrichment was reported as the odds ratio (OR), where I^A^ = # DEGs annotated with the GO term, I^B^ = # background genes annotated with the GO term, and OR = (I^A^/size of DEG list) / (I^B^/size of background list). Statistical significance was defined with a Fisher’s Exact Test.

### 2.4. Ingenuity Pathway Analyses

To analyze changes in biological states across the UC clinical subtypes, three sets of IPA Core Analyses were performed, followed by IPA Comparison Analysis. The IPA Comparison Analysis allows for side-by-side comparison of multiple Core Analyses, which facilitated the discovery of trends amongst the three datasets. Core Analyses were performed on the three datasets (i.e., left-sided colitis vs. HC, pancolitis vs. HC, and CAD vs. HC) to assess the canonical pathways, upstream regulators, molecular and cellular functions, and molecular interaction networks that were most likely to be perturbed based on the changes in gene expression. Within IPA, canonical pathways were built with reference to literature prior to DEG input and did not undergo structural changes upon DEG input. Instead, IPA computes a z-score that assesses the directionality within a gene set (i.e., the DEG input) to infer the activation state of each canonical pathway or molecular and cellular function. Upstream regulators were identified by the observed differential regulation of known downstream effector(s). The z-score determines the activation state of an upstream regulator by the regulation direction associated with the relationship from the upstream regulator to the effector(s). A negative z-score indicates inhibition, and a positive z-score indicates activation. Significance was calculated with the right-tailed Fisher’s Exact Test. Changes in activation state across UC subtypes were assessed with Comparison Analysis (sort method = trend + z-score). The correlation of activation state as the extent of disease progressed from left-sided colitis to pancolitis to CAD (the trend) was examined, and the findings were reported as trending towards activation or trending towards deactivation.

### 2.5. Mapping Molecular Interaction Networks

DEGs from the three datasets were mapped to their corresponding gene objects in the Ingenuity Knowledge Base (IKB), and those that interacted with other molecules in the IKB were designated focus molecules. Focus molecules were then assembled into networks by maximizing their interconnectedness with each other (relative to non-focus molecules with which they are connected to in the IKB). While non-focus molecules from the IKB may be used to merge smaller networks into a larger network, networks are scored based on the number of focus molecules they contain. Network size, the total number of focus molecules analyzed, and the total number of molecules in the IKB that could be included in the networks also contribute to the network scores. The score is a test of significance using hypergeometric distribution and is calculated with the right-tailed Fisher’s Exact Test (Score = - log (Fisher’s p-value; score ≥ 2 equals p ≤ 0.01). For this study, networks were limited to 70 molecules each, and a maximum of 25 networks per UC-subtype were constructed. Individual networks (child networks, count = 75) were overlapped with one another to create a single parent network by virtue of common network molecules between child pairings; this was done with the RVenn R package. Finally, the edge betweenness parameter for the core network was computed via the NetworkAnalyzer app, included in Cytoscape 3.8.0 (15).

### 2.6. Differential Correlations

A ggplot2-based R package, ggpubr (https://rpkgs.datanovia.com/ggpubr/), was used to investigate the relationship between the expression profiles of two genes. Specifically, function |stat_cor| was used to calculate the Pearson correlation coefficient. The size of the concentration ellipse in normal probability was left at the default 0.95, which translates to a 95% confidence interval.

## 3. RESULTS

### 3.1. Analysis of Genes Differentially Expressed in UC Disease Subtypes

The distribution of gene expression ratios (log-transformed) calculated between UC subtypes and healthy controls (HC) is presented in **Figure 1A**. For the left-sided colitis vs. HC comparison, 1,016 genes had log_2_FC greater than 0.75 (q < 0.001); of these DEGs, 614 were upregulated, and 402 were downregulated. Pancolitis returned 2,858 DEGs, half of which were upregulated. Colitis-associated dysplasia (CAD) returned 1,842 DEGs; 393 were upregulated, and 1,449 were downregulated.

**Figure 1.**
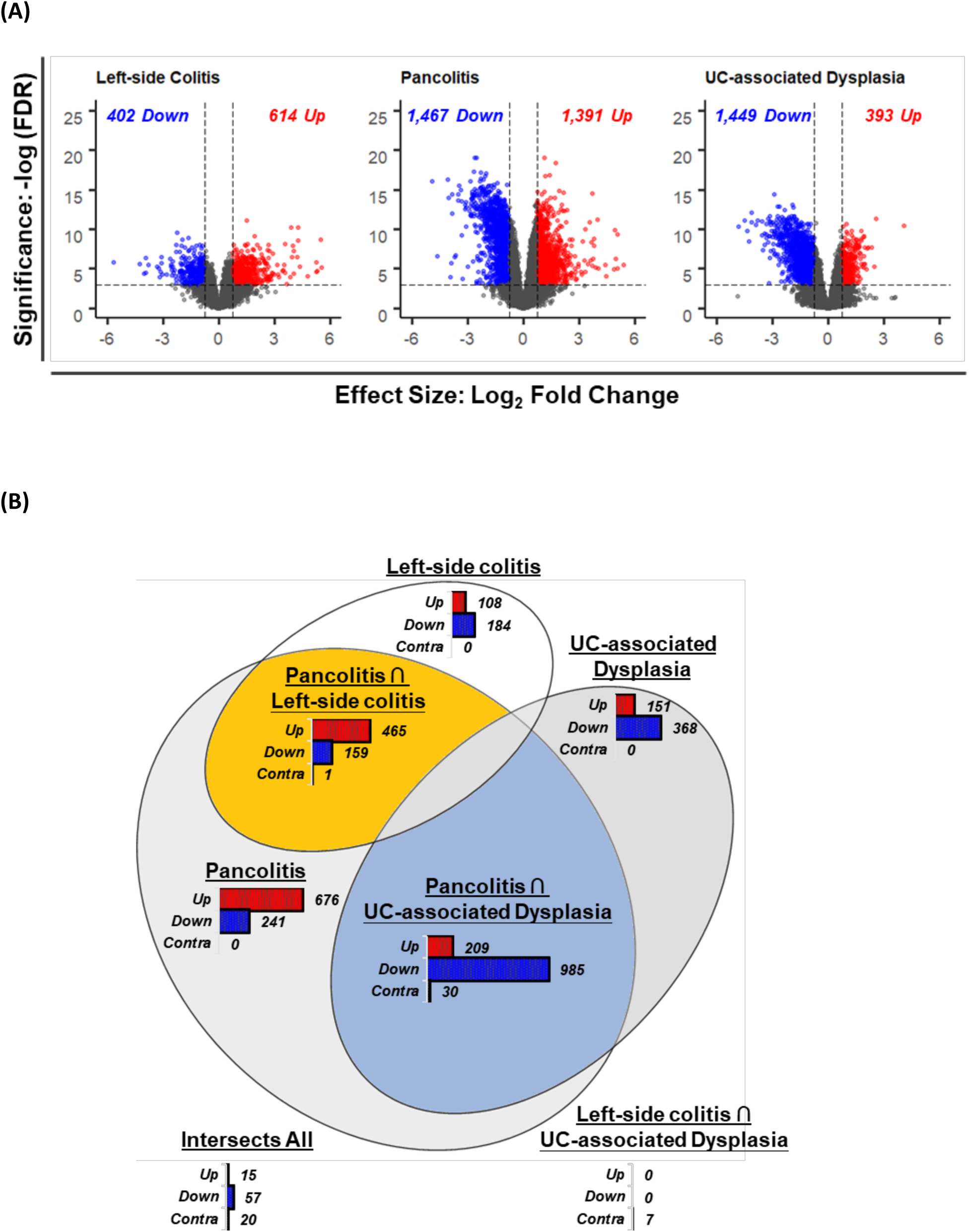
Differential expression of genes in UC disease subtypes. (**A**), differential gene expression of three UC clinical subtypes against healthy controls. Log2FC calculated with limma R for the GSE47908 dataset [n = 15 (healthy controls, HC), n = 20 (left-sided colitis), n = 19 (pancolitis), and n = 6 (colitis-associated dysplasia, CAD)]. The threshold set for log2FC was |0.75|. Significance was determined by FDR<0.001. (**B**), overlap of shared differentially expressed genes (DEGs) between UC subtypes. The proportional Venn diagram was generated with Bio Venn and then optimized with eulerAPE_3.0.0.jar.

For functional predictions, a Venn diagram was first used to identify genes with expression regulation observed in multiple UC disease subtypes. As presented in **Figure 1B**, pancolitis overlapped both left-sided colitis and CAD in terms of common DEGs. Of the 2,858 pancolitis DEGs, 651 shared similar expression regulation as left-sided colitis DEGs (465 common upregulation plus 159 common downregulation), and 1,194 shared similar expression regulations as CAD DEGs (209 common upregulation plus 985 common downregulation). This feature was not observed when comparing left-sided colitis and CAD; less than 2% of the input DEGs appeared at the intersection of left-sided colitis ∩ pancolitis ∩ CAD. This analysis suggests pancolitis exists as the middle ground of UC subtypes that bridges the progression of UC from left-sided colitis to CAD. Thus, the DEGs located at the intersection of pancolitis ∩ left-sided colitis or pancolitis ∩ CAD were selected for functional analysis.

To identify the potential biological processes associated with UC clinical progression, two sets of DEGs at the intersection of 1) pancolitis ∩ left-sided colitis, or 2) pancolitis ∩ CAD were processed separately with the topGO R package for functional enrichment analysis. These enrichment analyses included both up-regulated and down-regulated genes. The top 10 terms from the ‘Biological Process’ (BP) category of the GO enrichment analysis are presented in **Figure 2**. The DEGs at the intersection of pancolitis and left-sided colitis were enriched in inflammatory processes while the pancolitis ∩ CAD overlapping DEGs were enriched in actin-based processes. Specifically, the pancolitis ∩ left-sided colitis DEGs showed significant enrichment for the regulation of IFN-γ mediated signaling pathways (GO:0060334), the antimicrobial humoral immune response mediated by antimicrobial peptide (GO:0061844), and the acute-phase response (GO:0006953) with 8.6-, 6.4-, and 6.0-fold enrichments, respectively. Across the pancolitis ∩ CAD DEGs, microvillus assembly (GO:0030033), Golgi to plasma membrane transport (GO:0006893), and actin cytoskeleton reorganization (GO:0031532) were found significantly over-represented with 6.3-, 3.1-, and 3.0-fold enrichments, respectively. Overall, the GO enrichment analysis suggests that as UC progressed from left-sided colitis to pancolitis to CAD, the transcriptome pattern also shifted from being immune-centric to having cytoskeleton-dependence. Presumably, the regulation of actin cytoskeleton organization plays an important role in colitis-dysplasia progression.

**Figure 2.**
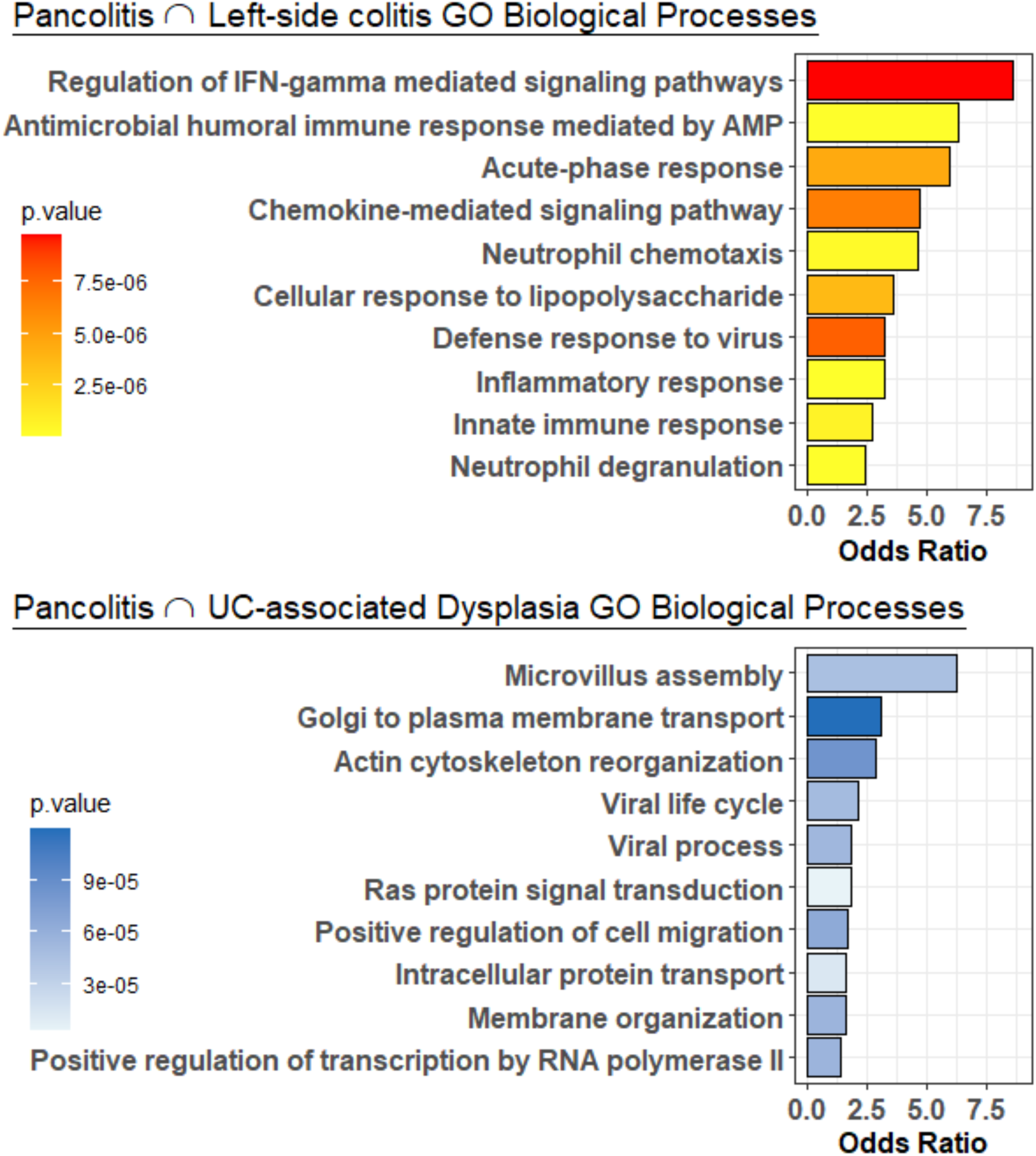
Gene ontology (GO) term enrichment analysis of differentially-expressed genes (DEGs) common among UC subtypes. DEGs at the intersection of pancolitis ∩ left-sided colitis or pancolitis ∩ CAD were subjected to GO term enrichment analysis for Biological Process categories. Enrichment was performed with topGO R (“elim” algorithm, “fisher” statistic). Statistical significance was defined with a Fisher’s Exact Test.

### 3.2. Association of Hippo Signaling Activation with Colitis-Dysplasia Progression

To identify canonical pathways that are most relevant to the observed shift in gene expression profile (**Figure 2**), a comparison analysis heat map for canonical pathways significantly altered by UC disease subtypes was constructed (**Figure 4**). This heatmap displays the trend of either activation or deactivation as the gene expression profile shifted in response to UC extent, that is, from left-sided colitis to pancolitis to CAD. The heatmap revealed Hippo signaling and Ephrin receptor signaling as the top canonical pathways that were progressively altered in concert with the pathogenic progression of UC. The Hippo signaling pathway displayed incremental activation, while the Ephrin receptor signaling pathway exhibited gradual deactivation as the UC extent broadened (**Figure 4**).

**Figure 3.**
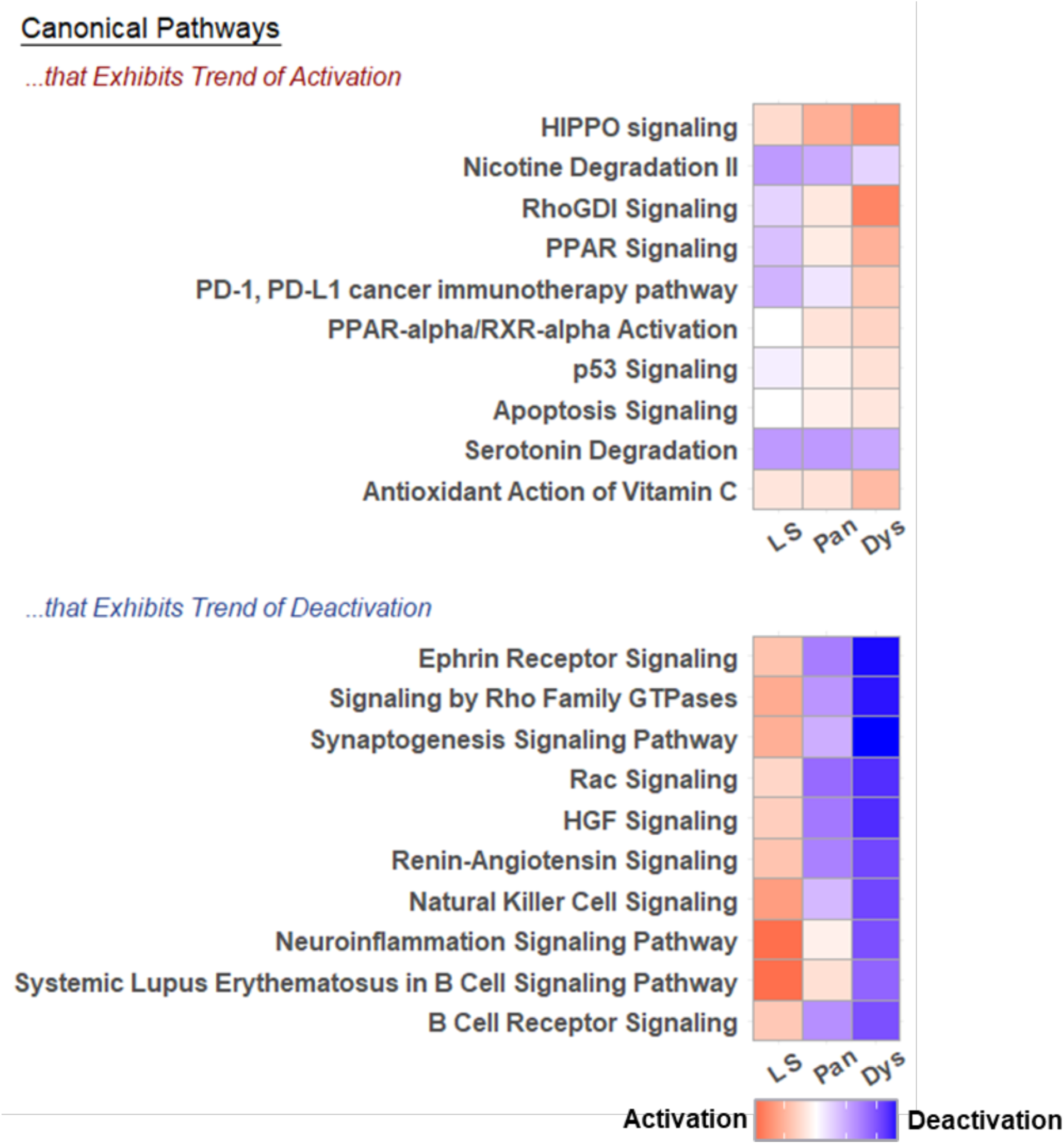
Ingenuity Pathway Analysis (IPA) of canonical pathways differentially deregulated within the UC subtypes. For each colitis subtype, the activation z-score calculated by IPA software predicts the activation (red) or inhibition (blue) potential for each canonical pathway. The pathways are sorted based on correlation of z-scores to extent of disease (left-sided colitis to pancolitis to CAD) with higher total scores across the colitis-subtypes ranked higher than those with lower total scores. LS: left-sided colitis, Pan: pancolitis, Dys: CAD.

**Figure 4.**
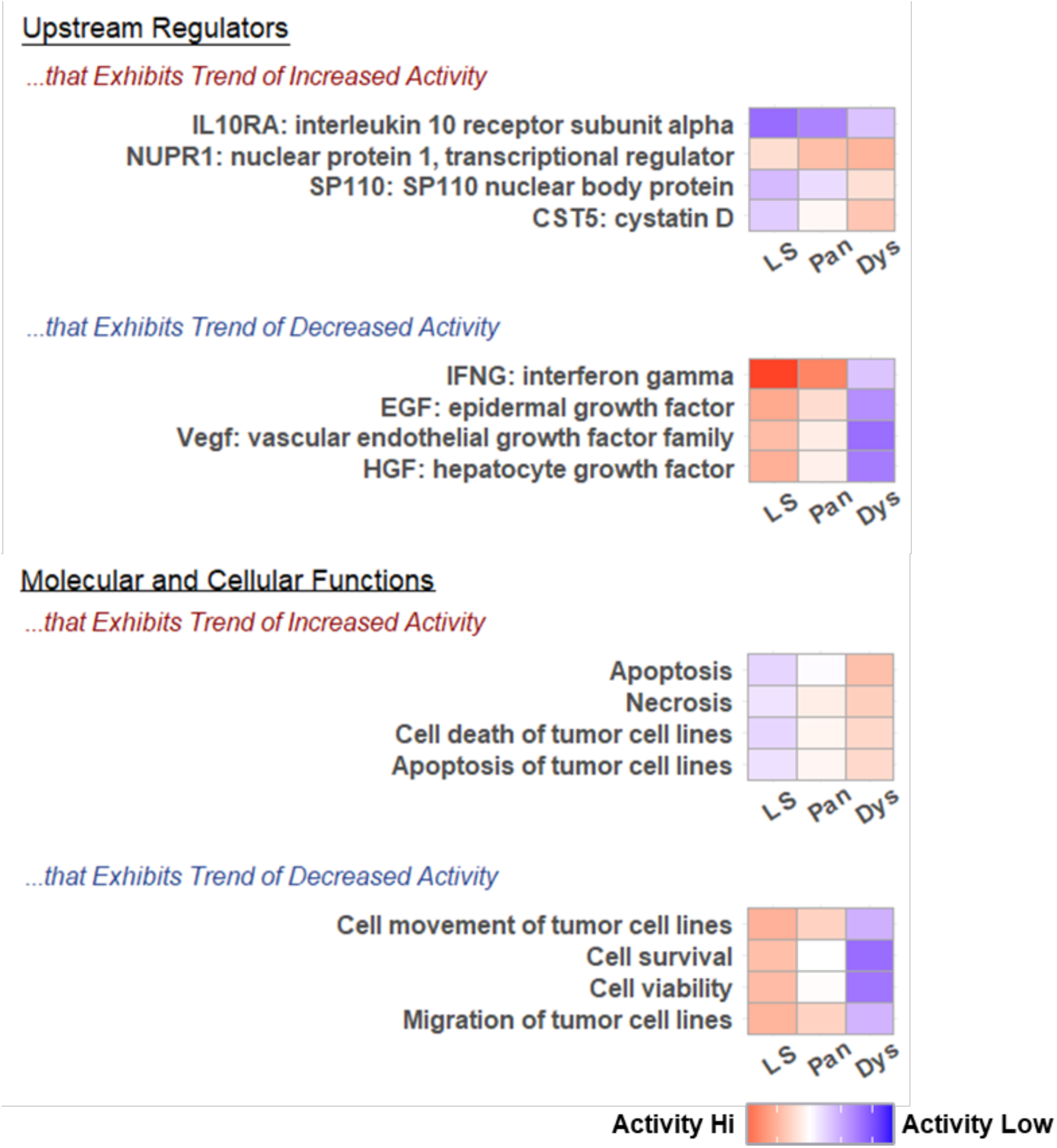
Ingenuity Pathway Analysis (IPA) of upstream regulators and functions differentially deregulated among UC subtypes. For each colitis subtype, the activation z-score calculated by IPA software predicts the activation (red) or inhibition (blue) potential for each regulator/function. The regulators/functions are sorted based on correlation of z-scores to extent of disease (left-sided colitis to pancolitis to CAD) with higher total scores across the colitis-subtypes ranked higher than those with lower total scores. LS: left-sided colitis, Pan: pancolitis, Dys: CAD.

Trends of activation or deactivation were also probed on upstream regulators and biological functions to obtain a basic view of the molecular mechanisms underlying the extent of colitis. In terms of upstream regulators, IL10RA exhibited a trend of increased activity whereas interferon (IFN)-γ exhibited a trend of decreased activity. Affected biological functions include apoptosis and cell movement, that underwent gradual activation and deactivation, respectively, as disease extended from left-sided colitis to pancolitis to CAD (**Figure 4**).

### 3.3. Actin Reorganization as a Potential Key Determinant for Colitis Progression

Using the limma R-derived DEGs, the IPA software generated 25 networks for each of the three UC subtypes. Molecular network intersections, assembled with the RVenn R package, revealed a single parent network that connected all 75 child networks via 757 intersections/edges (**Figure 5A**). Among the 75 child networks, approximately two-third (48/75) were composed of less than 60 focus molecules (<85% focus molecules). The 757 intersections/edges that connected the child networks each comprise one to 24 common molecule overlaps. Nevertheless, over half (57%) of the interactions/edges were enabled by just one common molecule, and 87% of the edges involved less than four common molecules (<5% overlap). To reduce noise from the parent network depicted in **Figure 5A**, molecular network intersection was compiled once more, but with child networks that were built upon 60 or more focus molecules (>85% focus molecules) and preserving connections that were maintained by four or more common molecules (>5% overlap). This produced one core network of networks that connected 21 child networks with 21 edges; One intersection segment, though strongly connecting pancolitis network 6 to CAD network 10 with 24 common molecules (34% overlap), stood apart from the core network of networks (**Figure 5B**), so was excluded from subsequent analysis.

**Figure 5.**
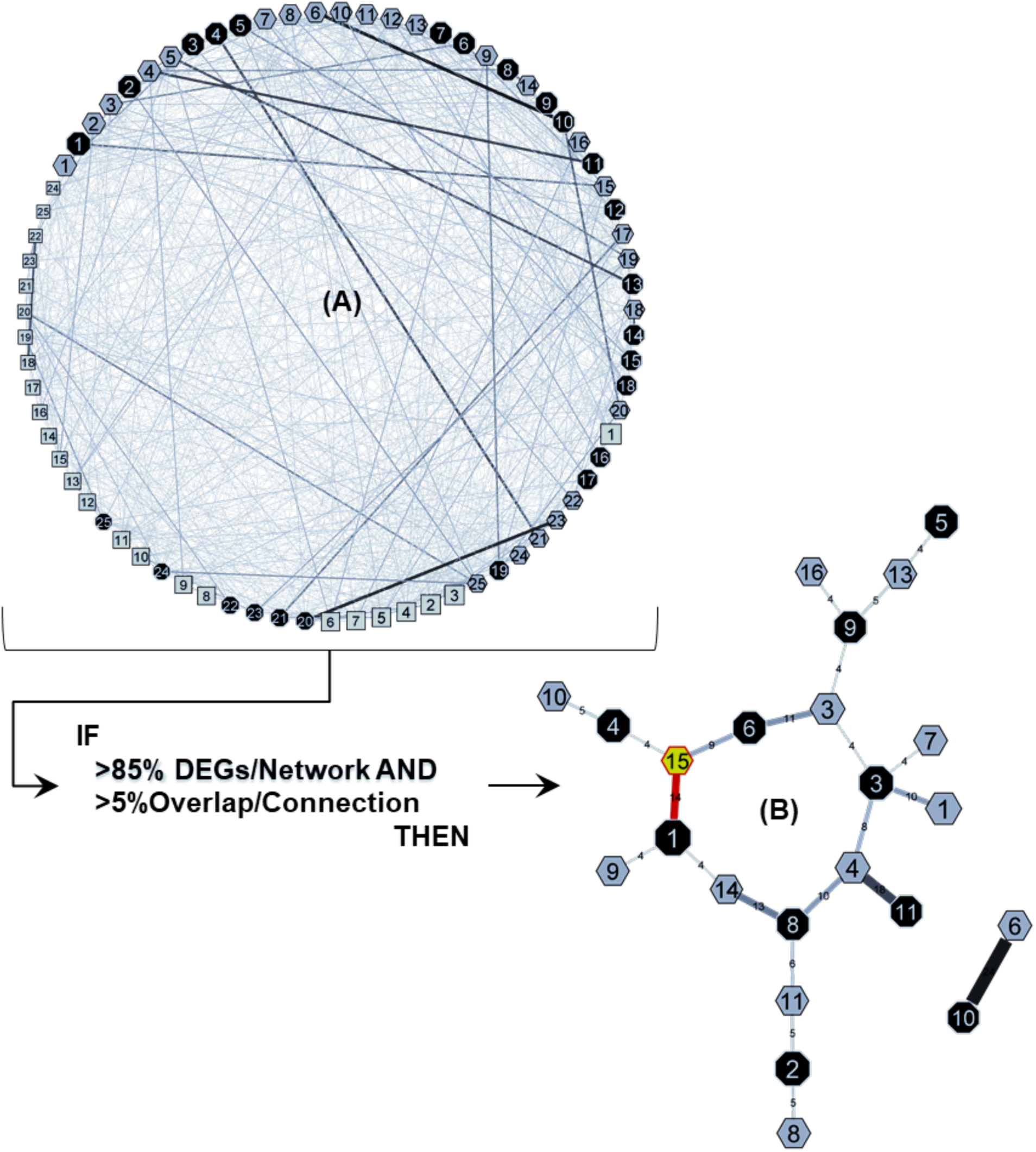
Intersections of UC clinical subtype molecular interactions. Molecular networks were constructed for each UC subtype. Octagons represent networks constructed with CAD DEGs, hexagons represent pancolitis networks, and rectangles indicate left-sided colitis networks. Shape size represents the relative number of DEGs used to build each network. The networks are connected to each other if there are any overlapping gene products, and line width represent the number of shared gene products. The serpentine network overlap (**A**) was detangled by focusing on networks that had >85% of DEG inputs and keeping edges that involved >5% overlap of gene products. This returned a core network of networks (**B**) that was heavily leveraged on the edge connecting CAD network 1 and pancolitis network 15 (red).

To pinpoint a network pairing that could be used to discern pathways or molecular processes bridging pancolitis to CAD, the ‘count of overlapping molecules’ and ‘value of edge betweenness’ measures were utilized for edge evaluation. Edge betweenness reflects the amount of control that an edge exerts over the interactions of other child networks in the parent network. The edge betweenness of e=(v,w) is defined as the number of shortest paths between two nodes that go through e divided by the total number of shortest paths that go from the two nodes (16,17). Edge betweenness does not consider the number of overlapping molecules that contribute to the intrinsic strength of each edge. Thus, additional consideration for this attribute was completed *post hoc*. As shown in **Figure 5B** and **Table 1**, the edge connecting pancolitis network 15 to CAD network 1 stood out from the remaining network associations by virtue of its large number of overlapping molecules (20% overlap) and high value of edge betweenness. Taken together, the findings suggest that the removal of this edge may affect interactions between the remaining networks within network. Thus, the overlap between pancolitis network 15 and CAD network 1 was selected for further detailed examination.

**TABLE 1.** Edge attributes of the core multiplex bridging pancolitis to CAD.

| Edge | No. of Overlapping Molecules | Edge-Betweenness |
| --- | --- | --- |
| CAD.1 (Overlap) Pan.15 | 14 | 89 |
| CAD.8 (Overlap) Pan.14 | 13 | 87 |
| CAD.6 (Overlap) Pan.3 | 11 | 109 |
| CAD.8 (Overlap) Pan.4 | 10 | 119 |
| CAD.6 (Overlap) Pan.15 | 9 | 101 |
| CAD.3 (Overlap) Pan.4 | 8 | 127 |
| CAD.8 (Overlap) Pan.11 | 6 | 108 |
| CAD.3 (Overlap) Pan.3 | 4 | 139 |
| CAD.8 (Overlap) Pan.3 | 4 | 136 |
| CAD.1 (Overlap) Pan.14 | 4 | 81 |

A network of networks (i.e., multiplex) approach was used to study the joint organization of transcriptional networks defining pancolitis (Pan) and colitis-associated dysplasia (CAD). The edge connecting CAD network 1 and pancolitis network 15 was prioritized based on the number of overlapping molecules and degree of betweenness. The Edge-Betweenness was calculated with the Cytoscape app NetworkAnalyzer and reflects the amount of control that an edge exerts over the interactions of other nodes in the network. This measure does not consider the number of overlapping molecules that contributes to the intrinsic strength of each edge. Consideration for the strength attribute was computed *post hoc*.

The intersection pancolitis 15 ∩ CAD 1 was formed by 14 gene products, which, when analyzed for GO term enrichment, was predominantly composed of actin-based processes (**Figure 6**). Specifically, the DEGs at this intersection showed significant enrichment for the ruffle organization (GO:0031529), the regulation of actin filament polymerization (GO:0030833), and the myofibril assembly (GO:0030239) with 66-, 33-, and 33-fold enrichments, respectively.

**Figure 6.**
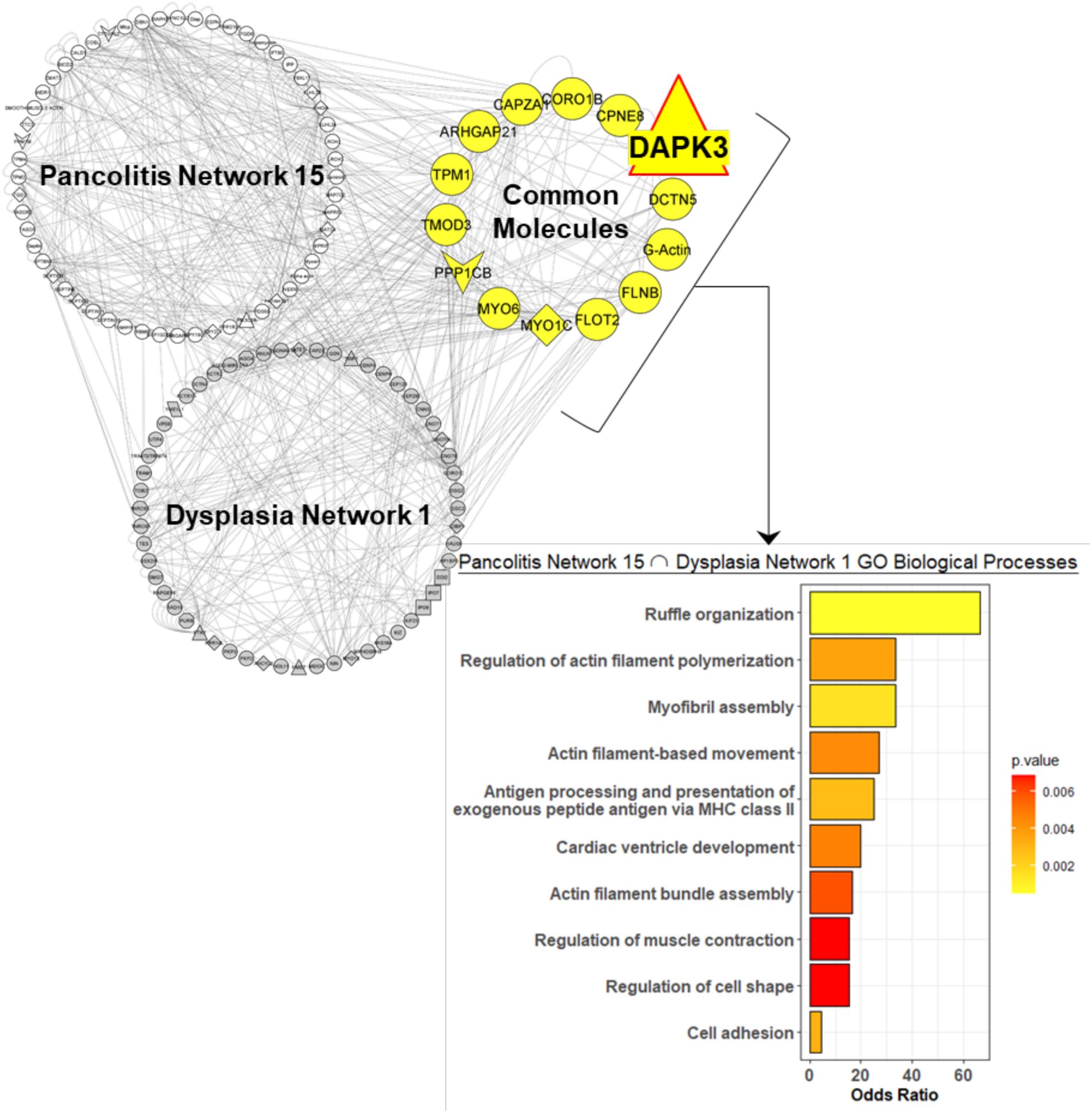
Gene ontology (GO) term enrichment analysis of the pancolitis network 15 and CAD network 1 intersection. DEGs at the intersection of pancolitis network 15 ∩ CAD (colitis-associated dysplasia) network 1 were subjected to GO term enrichment analysis for Biological Process categories. Network constructed by IPA software. Network intersection was compiled with RVenn R. Edge-betweenness was computed by the Cytoscape app NetworkAnalyzer. Enrichment performed with topGO R. Statistical significance was defined with a Fisher’s Exact Test.

### 3.4. Implication of DAPK3 as a Key Factor in the Colitis-Dysplasia Progression

Further dissection of pancolitis 15 and CAD 1 revealed two signal transduction gene products that reside in the overlap of these two networks (**Table 2**). Namely, death-associated protein kinase 3 (DAPK3) as the sole protein kinase, and protein phosphatase PP1β catalytic subunit (PPP1CB) as the sole protein phosphatase. The remaining 12 gene products were categorized as either mechanochemical enzymes (i.e., motors) or scaffolding proteins involving the cytoskeleton. Both DAPK3 and PPP1CB were downregulated in pancolitis (log2FC: -1.08, DAPK3; -1.56, PPP1CB) and CAD (log2FC: -0.98, DAPK3; -1.52, PPP1CB) but showed no deregulation in left-sided colitis. Presumably, DAPK3 and PPP1CB act as key factors regulating the altered molecular pathways that bridge pancolitis to CAD.

**TABLE 2.** Overlapping gene products in the pancolitis 15 ∩ CAD network 1 intersection.

| Gene | Protein | Molecular Function | Expression FC |  |  |
| --- | --- | --- | --- | --- | --- |
|  |  |  | L | P | D |
| *DAPK3* | Death-associated protein kinase 3 | Kinase | -0.15 | -1.08<sup>§</sup> | -0.98<sup>§</sup> |
| *PPP1CB* | Protein phosphatase PP1β subunit | Phosphatase | -0.12 | -1.56<sup>§</sup> | -1.52<sup>§</sup> |
| *MYO1C* | Unconventional myosin IC | Motor | -0.18 | 0.77* | 0.99* |
| *MYO6* | Unconventional myosin VI | Motor | -0.31 | -0.88<sup>§</sup> | -1.36<sup>§</sup> |
| *ARHGAP21* | Rho GTPase-activating protein 21 | GTPase | -0.48 | -0.98<sup>§</sup> | -0.97<sup>§</sup> |
| *CAPZA1* | F-actin-capping protein α1 | Actin Binding | 0.18 | -1.89<sup>§</sup> | -2.21<sup>§</sup> |
| *CORO1B* | Coronin 1B | Actin Binding | -0.08 | 0.88* | 0.86* |
| *CPNE8* | Copine 8 | Membrane Binding | -0.34 | -1.55<sup>§</sup> | -1.93<sup>§</sup> |
| *DCTN5* | Dynactin 5 | Scaffold | -0.20 | -0.80<sup>§</sup> | -1.64<sup>§</sup> |
| *FLNB* | Filamin B | Actin Binding | -0.49 | -0.76<sup>§</sup> | -1.20<sup>§</sup> |
| *FLOT2* | Flotillin 2 | Scaffold | 0.17 | -0.78<sup>§</sup> | -0.95<sup>§</sup> |
| *TMOD3* | Tropomodulin 3 | Actin Binding | -0.22 | -1.36<sup>§</sup> | -2.15<sup>§</sup> |
| *TPM1* | Tropomyosin α1 | Actin Binding | -0.61 | -0.86<sup>§</sup> | 1.01* |
| --- | Monomeric (G) actin | Structural | Not a focus molecule |  |  |

FC = log<sub>2</sub> fold-change, L = left-sided colitis, P = pancolitis, and D = colitis-associated dysplasia. A gene was deemed dysregulated when $|\log_2\text{FC}| > 0.75$ and $\text{FDR} < 0.001$ ; \* - significant upregulation, § - significant downregulation.

### 3.5. Differential Correlation of DAPK3-YAP with UC Disease Progression

Given the association of Hippo signaling activation with UC extent (**Figure 3**), and the implication of *DAPK3* and *PPP1CB* as key factors in colitis-dysplasia progression (**Figure 6**), differential correlation analyses for the pairings *DAPK3-YAP* and *PPP1CB-YAP* were conducted to assess potential differences in gene-gene regulatory relationships among the UC disease subtypes. As shown in **Figure 7A**, the *DAPK3-YAP* correlation in healthy controls was negative but flipped to positive in left-sided colitis. Moreover, as the UC disease extent progressed from left-sided colitis to pancolitis and then on to CAD, the *DAPK3-YAP* correlation grew progressively more positive. The general direction of differential correlation for the *PPP1CB-YAP* pairing mirrors that of the *DAPK3-YAP* pairing (**Figure 7B**). However, the differential correlation with UC disease extent was less apparent for the *PPP1CB-YAP* pairing. This result suggests that changes in the potential regulatory relationship between DAPK3 and YAP, conditioned on UC disease extent, may contribute to disease progression.

**Figure 7.**
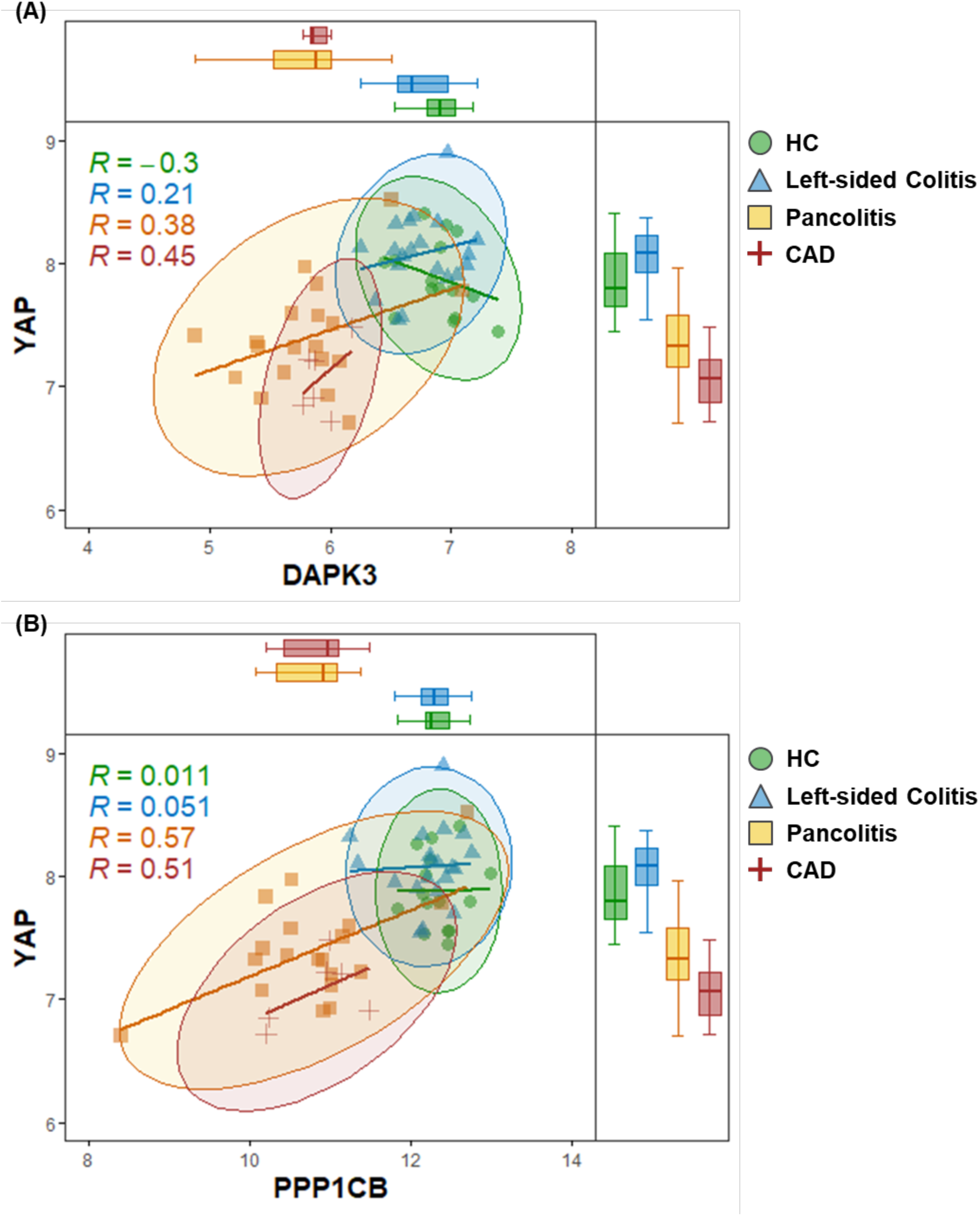
Differential correlation of DAPK3-YAP and PPP1CB-YAP relationships. The main panels display the relationship between the log-transformed expression profiles of (**A**) *DAPK3* and *YAP* or (**B**) *PPP1CB* and *YAP*. These scatterplots are overlayed with confidence ellipses of covariance, constructed for the 95% CI. In the marginal box plots of log-transformed gene expression, the lower whisker represents the lowest datum still within 1.5 interquartile range (IQR) of the lower quartile, and the upper whisker represents the highest datum still within 1.5 IQR of the upper quartile.

## 4. DISCUSSION

In UC, prolonged disease duration and extensive intestinal involvement (i.e., pancolitis) are associated with an increased risk for colorectal cancer (7). The progression of carcinogenesis in UC is thought to be driven by chronic inflammation and proceeds in a stepwise manner through a colitis-dysplasia-carcinoma sequence (18-21). As the area affected by UC grows so too does the inflammatory load which could in turn accelerate dysplasia and CAC tumorigenesis (20). While the impact that inflammation can have on colon carcinogenesis should not be discounted, evidence also suggests that non-inflammatory factors play a role in mediating the colitis-dysplasia-carcinoma progression (5,22,23). Among patients or animals with similar inflammatory status, some develop CAC while others do not (5). Additionally, whole-exome sequence analysis of IBD-associated colorectal cancer showed that, apart from rare cases of mutations in DNA proofreading or repair pathways, supra-IBD inflammation alone does not compel greater mutation rates when compared with sporadic CRC of non-inflammatory origin (23). This also supports a role for non-inflammatory factors in mediating colitis-dysplasia-carcinoma progression. As an example, Arthur and colleagues found that while azoxymethane (AOM)-treated *Il10*-/-mice infected with colitogenic *E. coli* NC101 or *E. faecalis* OG1RF exhibited a similar degree of colitis and comparable levels of immune infiltrate, tumors were observed in only 10% of *E. faecalis* infected mice whereas 80% of *E. coli* infected mice showed tumor development. The authors then associated the polyketide synthase (*pks*) genotoxic island, found in *E. coli* NC101 but not *E. faecalis* OG1RF, with the DNA damage and subsequent tumorigenesis in their *E. coli* infected mice (24). Moreover, gene mutation and gene expression analyses have linked cytoskeleton remodeling to colitis-dysplasia-carcinoma progression (23,25,26). Utilizing the Bjerrum dataset, our investigation verified pancolitis as a conduit for UC advancement from left-sided colitis to CAD and confirmed dysregulation of actin reorganization as a key determinant for the progression of UC from non-dysplastic to dysplastic UC.

Out of the Bjerrum dataset, 651 and 1,194 parallel dysregulated transcripts were identified at the intersection of pancolitis ∩ left-sided colitis and pancolitis ∩ CAD, respectively. GO term enrichment analysis revealed that the parallelisms of pancolitis with CAD were rooted in dysregulation of actin-based processes, whereas the similarities between pancolitis and left-sided colitis were rooted in dysregulation of inflammatory processes. This finding substantiates results of a gene expression analysis of patients with CAC presented by Kanaan and colleagues, identifying actin cytoskeleton organization as the most significantly disrupted process in UC progression (25). Protein abundance analysis of UC progressors (UC patients with CAD or CAC) versus UC non-progressors has also identified enrichment of dysregulated cytoskeletal proteins in UC progressors (27). Drawbacks in the two aforementioned studies include the evaluation of UC progression in a binary fashion (presence or absence of CAD and/or CAC) as well as the small sample sizes (i.e., three unique patients in the study conducted by Kanaan and colleagues (25), and 15 unique patients in the study conducted by May and colleagues (27)). To provide insight into the molecular events associated with the stepwise progression of UC, it will be necessary to demonstrate that the dysregulations observed in CAD or CAC were issued forth from some form of non-dysplastic UC.

Signal transduction pathways regulate many cellular functions that were found altered in UC carcinogenesis, such as proliferation, growth, differentiation, metabolism, and survival. Molecular investigations have previously associated STAT3, Wnt, TGF-β, and TLR4/NFκB signaling with the pathogenesis of colon carcinogenesis (28-32). However, the pathways most relevant to the gene expression changes observed during the early progression of UC, before any histological evidence of dysplasia/carcinoma, remain unknown. In this regard, Hippo signaling and Ephrin receptor signaling were uncovered as the top canonical pathways that were progressively altered in concert with the extent of UC progression (**Figure 3**). The Hippo pathway is a fundamental signaling cascade that negatively regulates the activity of YAP/TAZ to coordinate cell proliferation, apoptosis, and cell movement; as such, it is essential for tissue homeostasis, repair, and regeneration (33). Importantly, YAP/TAZ-mediated cell proliferation in epithelial monolayers is controlled by a cytoskeletal checkpoint, which in turn, is monitored by actin-processing factors. The Ephrin pathway also controls intestinal homeostasis through cell proliferation and cell movement additionally to cell attachment and repulsion (34-36). However, deciphering functional outcomes by Ephrin pathway activation is circuitous due to the redundancy and idiosyncrasy of this pathway (37). The Ephrin receptors (Eph) comprise the largest family of receptor tyrosine kinases (RTK). But, unlike most RTKs for which ligands are generally soluble, the cognate ligands of Eph receptors, the ephrins, are also membrane bound. This aspect of the Eph-ephrin receptor-ligand pairing consequently induces bidirectional signaling, where signaling through Eph is termed forward signaling and through ephrin is termed reverse signaling. Furthermore, there is a plethora of Ephs and ephrins (i.e., 14 Ephs, 8 ephrins) with promiscuous pairing options. Finally, Eph/ephrin also exhibits cis-interactions to inhibit forwarding signaling. Ephrin is therefore a more convoluted pathway to render, relative to the Hippo pathway.

The Hippo pathway was previously identified as a key factor for the compensatory regeneration of IECs in response to tissue injury using the dextran sodium sulphate (DSS)-induced colitis mouse model (38). The recent emergence of YAP as a potential regulator of intestinal diseases involves elements beyond the canonical Hippo pathway. For one, YAP can be sequestered at adherens junctions (AJs) via interactions with α-catenin (39), the abundance of which is significantly altered in active UC (40). Secondly, nuclear translocation of YAP may be brought about via stimulation of gp130-associated Src family kinase Yes (41). Finally, YAP was found to be a crucial pivot point of cellular reprograming during intestinal epithelial repair, coupling epithelial restitution to the proliferative phase of regeneration by way of FAK-Src signaling (42). In view of YAP as a mechanosensor and mechanotranducer amidst epithelial regeneration of injured tissue, a comprehensive understanding of the interplay between YAP and the actin cytoskeleton is needed to make rational selections of therapeutic targets for patients at high risk of UC neoplastic progression.

The identification of DAPK3 as a potential key factor in UC progression is particularly interesting, and the role for DAPK3 in UC pathogenesis was unknown prior to this study. However, DAPK1, the closely-related family member and upstream regulator of DAPK3, was previously associated with UC severity (43) and gastrointestinal cancer pathogenesis (44). In addition, pharmacological inhibition of DAPK1 was reported to augment susceptibility to DSS-induced colitis in mice, with concomitant increase in bacterial translocation ascribed to epithelial barrier defects (45). It is regrettable that the small molecule inhibitor of DAPK1 (i.e., DAPK6 or DI) applied in the study of tunicamycin (TM)-induced, ER-stress-dependent reduction of bacterial translocation, potently cross-inhibits DAPK3 (IC_50_=225 nM (46)) and Rho-associated coil-coiled kinase (ROCK, K_i_=132 nM (47)). Although the Lopes study included siRNA knockdown experiments to independently validate DAPK1 signaling involvement, the potential impact of concurrent DAPK3 and ROCK inhibition brought forth with the DAPK6 inhibitor was not examined. Previously, Ito and colleagues showed that ROCK activity increased in response to TM, and that treatment with Y27632 (a ROCK inhibitor: K_i_=220 nM for ROCK1, K_i_=300 nM for ROCK2 (48,49)) completely reversed TM-induced ER-stress responses in the J774 macrophage cell line (50). Moreover, the involvement of DAPK3 in ER-stress response was also demonstrated in human aortic vascular smooth muscle cells, where shRNA-mediated silencing of *DAPK3* ablated the calcifying-media induced increase of CCAAT-enhancer-binding protein homologous protein (CHOP), a multifunctional transcription factor in ER-stress response (51). In the same study, treatment with DAPK6 attenuated vascular calcification in rats, alongside a significant reduction in CHOP protein abundance in the aorta. It may be beneficial to learn whether DAPK3 and/or ROCK alter ER-stress-dependent autophagy in the context of DSS-induced colitis in mice.

The probability that DAPK3 plays a role in the progression of the pathological changes of UC is notable. To substantiate the connection of DAPK3 to UC progression, the difference in correlation between *DAPK3* and *YAP* was studied in all UC clinical subtypes plus healthy control samples. Differential co-expression operates on the level of gene pairs and is used as an alternative approach to identify disease-related genes (52,53). Results from the differential co-expression analyses demonstrate the correspondence of *DAPK3*-*YAP* correlation with UC extent. This suggests that changes in the potential regulatory relationship between DAPK3 and YAP, conditioned on UC extent, may contribute to UC progression. Still, the driver(s) behind the differential *DAPK3*–*YAP* co-expression pattern is unclear. Better understanding of the DAPK3-YAP relationship may enable the discovery of targeted therapy for the prevention of UC neoplastic progression.

### Study Limitations

The progression of UC to CAD occurs through multiple mechanisms involving various cell types. The present analyses were completed on transcriptional profiles generated from mucosal biopsies (8), and genes may demonstrate diverse functions across different cell types. Hence, the gene sets identified from the averaged dataset will require re-examination in a cell type-specific way (e.g., single cell RNA-Seq) to precisely identify the susceptible cell types and convergent pathways among different cells.

## Data Availability

The GSE47908 transcriptional microarray dataset is available to the public via the Gene Expression Omnibus.

https://www.ncbi.nlm.nih.gov/geo/query/acc.cgi?acc=GSE47908

## 5. ACKNOWLEDGEMENTS

This work was supported by a research grant from the Canadian Institutes of Health Research (MOP#97931 to J.A.M.). H.-M.C. was recipient of CIHR Fredrick Banting and Charles Best Canada and Alberta Graduate Excellence Scholarships.

## 6. AUTHOR CONTRIBUTIONS

H.-M.C. completed the data analysis, prepared figures and wrote the manuscript. J.A.M. conceived and coordinated the study, wrote the manuscript, supervised trainees and provided intellectual contributions to the project. All authors reviewed the results and approved the final version of the manuscript.

## 7. AUTHORS’ DECLARATION OF INTERESTS STATEMENT

J.A.M. is cofounder and has an equity position in Arch Biopartners Inc. All other authors declare no conflicts of interest.

## Notes

### Competing Interest Statement

The authors have declared no competing interest.

## REFERENCES

1. Ordas, I., Eckmann, L., Talamini, M., Baumgart, D. C., and Sandborn, W. J. (2012) Ulcerative colitis. Lancet 380, 1606–1619

2. Porter, R. J., Kalla, R., and Ho, G. T. (2020) Ulcerative colitis: Recent advances in the understanding of disease pathogenesis. F1000Res 9

3. Younis, N., Zarif, R., and Mahfouz, R. (2020) Inflammatory bowel disease: between genetics and microbiota. Mol Biol Rep 47, 3053–3063

4. Jostins, L., Ripke, S., Weersma, R. K., Duerr, R. H., McGovern, D. P., Hui, K. Y., Lee, J. C., Schumm, L. P., Sharma, Y., Anderson, C. A., Essers, J., Mitrovic, M., Ning, K., Cleynen, I., Theatre, E., Spain, S. L., Raychaudhuri, S., Goyette, P., Wei, Z., Abraham, C., Achkar, J. P., Ahmad, T., Amininejad, L., Ananthakrishnan, A. N., Andersen, V., Andrews, J. M., Baidoo, L., Balschun, T., Bampton, P. A., Bitton, A., Boucher, G., Brand, S., Buning, C., Cohain, A., Cichon, S., D’Amato, M., De Jong, D., Devaney, K. L., Dubinsky, M., Edwards, C., Ellinghaus, D., Ferguson, L. R., Franchimont, D., Fransen, K., Gearry, R., Georges, M., Gieger, C., Glas, J., Haritunians, T., Hart, A., Hawkey, C., Hedl, M., Hu, X., Karlsen, T. H., Kupcinskas, L., Kugathasan, S., Latiano, A., Laukens, D., Lawrance, I. C., Lees, C. W., Louis, E., Mahy, G., Mansfield, J., Morgan, A. R., Mowat, C., Newman, W., Palmieri, O., Ponsioen, C. Y., Potocnik, U., Prescott, N. J., Regueiro, M., Rotter, J. I., Russell, R. K., Sanderson, J. D., Sans, M., Satsangi, J., Schreiber, S., Simms, L. A., Sventoraityte, J., Targan, S. R., Taylor, K. D., Tremelling, M., Verspaget, H. W., De Vos, M., Wijmenga, C., Wilson, D. C., Winkelmann, J., Xavier, R. J., Zeissig, S., Zhang, B., Zhang, C. K., Zhao, H., International, I. B. D. G. C., Silverberg, M. S., Annese, V., Hakonarson, H., Brant, S. R., Radford-Smith, G., Mathew, C. G., Rioux, J. D., Schadt, E. E., Daly, M. J., Franke, A., Parkes, M., Vermeire, S., Barrett, J. C., and Cho, J. H. (2012) Host-microbe interactions have shaped the genetic architecture of inflammatory bowel disease. Nature 491, 119–124

5. Van Der Kraak, L., Gros, P., and Beauchemin, N. (2015) Colitis-associated colon cancer: Is it in your genes? World J Gastroenterol 21, 11688–11699

6. Baker, A. M., Cross, W., Curtius, K., Al Bakir, I., Choi, C. R., Davis, H. L., Temko, D., Biswas, S., Martinez, P., Williams, M. J., Lindsay, J. O., Feakins, R., Vega, R., Hayes, S. J., Tomlinson, I. P. M., McDonald, S. A. C., Moorghen, M., Silver, A., East, J. E., Wright, N. A., Wang, L. M., Rodriguez-Justo, M., Jansen, M., Hart, A. L., Leedham, S. J., and Graham, T. A. (2019) Evolutionary history of human colitis-associated colorectal cancer. Gut 68, 985–995

7. Zhou, Q., Shen, Z. F., Wu, B. S., Xu, C. B., He, Z. Q., Chen, T., Shang, H. T., Xie, C. F., Huang, S. Y., Chen, Y. G., Chen, H. B., and Han, S. T. (2019) Risk of Colorectal Cancer in Ulcerative Colitis Patients: A Systematic Review and Meta-Analysis. Gastroenterol Res Pract 2019, 5363261

8. Bjerrum, J. T., Nielsen, O. H., Riis, L. B., Pittet, V., Mueller, C., Rogler, G., and Olsen, J. (2014) Transcriptional analysis of left-sided colitis, pancolitis, and ulcerative colitis-associated dysplasia. Inflamm Bowel Dis 20, 2340–2352

9. Davis, S., and Meltzer, P. S. (2007) GEOquery: a bridge between the Gene Expression Omnibus (GEO) and BioConductor. Bioinformatics 23, 1846–1847

10. Ritchie, M. E., Phipson, B., Wu, D., Hu, Y., Law, C. W., Shi, W., and Smyth, G. K. (2015) limma powers differential expression analyses for RNA-sequencing and microarray studies. Nucleic Acids Res 43, e47

11. Kramer, A., Green, J., Pollard, J., Jr., and Tugendreich, S. (2014) Causal analysis approaches in Ingenuity Pathway Analysis. Bioinformatics 30, 523–530

12. Blighe, K., Rana, S., and Lewis, M. (2021) EnhancedVolcano: Publication-ready volcano plots with enhanced colouring and labeling. R package v1.10.0

13. Alexa, A., and Rahnenfuhrer, J. (2021) topGO: Enrichment Analysis for Gene Ontology. R package v 2.44.0

14. Carlson, M. (2019) org.Hs.eg.db: Genome wide annotation for Human. R package v3.8.2

15. Otasek, D., Morris, J. H., Boucas, J., Pico, A. R., and Demchak, B. (2019) Cytoscape Automation: empowering workflow-based network analysis. Genome Biol 20, 185

16. Newman, M. E., and Girvan, M. (2004) Finding and evaluating community structure in networks. Phys Rev E Stat Nonlin Soft Matter Phys 69, 026113

17. Yoon, J., Blumer, A., and Lee, K. (2006) An algorithm for modularity analysis of directed and weighted biological networks based on edge-betweenness centrality. Bioinformatics 22, 3106–3108

18. Itzkowitz, S. H., and Yio, X. (2004) Inflammation and cancer IV. Colorectal cancer in inflammatory bowel disease: the role of inflammation. Am J Physiol Gastrointest Liver Physiol 287, G7–17

19. Dulai, P. S., Sandborn, W. J., and Gupta, S. (2016) Colorectal Cancer and Dysplasia in Inflammatory Bowel Disease: A Review of Disease Epidemiology, Pathophysiology, and Management. Cancer Prev Res (Phila) 9, 887–894

20. Grivennikov, S. I. (2013) Inflammation and colorectal cancer: colitis-associated neoplasia. Semin Immunopathol 35, 229–244

21. Low, D., Mino-Kenudson, M., and Mizoguchi, E. (2014) Recent advancement in understanding colitis-associated tumorigenesis. Inflamm Bowel Dis 20, 2115–2123

22. Grivennikov, S. I., and Cominelli, F. (2016) Colitis-Associated and Sporadic Colon Cancers: Different Diseases, Different Mutations? Gastroenterology 150, 808–810

23. Robles, A. I., Traverso, G., Zhang, M., Roberts, N. J., Khan, M. A., Joseph, C., Lauwers, G. Y., Selaru, F. M., Popoli, M., Pittman, M. E., Ke, X., Hruban, R. H., Meltzer, S. J., Kinzler, K. W., Vogelstein, B., Harris, C. C., and Papadopoulos, N. (2016) Whole-Exome Sequencing Analyses of Inflammatory Bowel Disease-Associated Colorectal Cancers. Gastroenterology 150, 931–943

24. Arthur, J. C., Perez-Chanona, E., Muhlbauer, M., Tomkovich, S., Uronis, J. M., Fan, T. J., Campbell, B. J., Abujamel, T., Dogan, B., Rogers, A. B., Rhodes, J. M., Stintzi, A., Simpson, K. W., Hansen, J. J., Keku, T. O., Fodor, A. A., and Jobin, C. (2012) Intestinal inflammation targets cancer-inducing activity of the microbiota. Science 338, 120–123

25. Kanaan, Z., Qadan, M., Eichenberger, M. R., and Galandiuk, S. (2010) The actin-cytoskeleton pathway and its potential role in inflammatory bowel disease-associated human colorectal cancer. Genet Test Mol Biomarkers 14, 347–353

26. Majumdar, D., Tiernan, J. P., Lobo, A. J., Evans, C. A., and Corfe, B. M. (2012) Keratins in colorectal epithelial function and disease. Int J Exp Pathol 93, 305–318

27. May, D., Pan, S., Crispin, D. A., Lai, K., Bronner, M. P., Hogan, J., Hockenbery, D. M., McIntosh, M., Brentnall, T. A., and Chen, R. (2011) Investigating neoplastic progression of ulcerative colitis with label-free comparative proteomics. J Proteome Res 10, 200–209

28. Chandrasinghe, P., Cereser, B., Moorghen, M., Al Bakir, I., Tabassum, N., Hart, A., Stebbing, J., and Warusavitarne, J. (2018) Role of SMAD proteins in colitis-associated cancer: from known to the unknown. Oncogene 37, 1–7

29. Claessen, M. M., Schipper, M. E., Oldenburg, B., Siersema, P. D., Offerhaus, G. J., and Vleggaar, F. P. (2010) WNT-pathway activation in IBD-associated colorectal carcinogenesis: potential biomarkers for colonic surveillance. Cell Oncol 32, 303–310

30. Muller, M., Hansmannel, F., Arnone, D., Choukour, M., Ndiaye, N. C., Kokten, T., Houlgatte, R., and Peyrin-Biroulet, L. (2020) Genomic and molecular alterations in human inflammatory bowel disease-associated colorectal cancer. United European Gastroenterol J 8, 675–684

31. Shenoy, A. K., Fisher, R. C., Butterworth, E. A., Pi, L., Chang, L. J., Appelman, H. D., Chang, M., Scott, E. W., and Huang, E. H. (2012) Transition from colitis to cancer: high Wnt activity sustains the tumor-initiating potential of colon cancer stem cell precursors. Cancer Res 72, 5091–5100

32. Tang, A., Li, N., Li, X., Yang, H., Wang, W., Zhang, L., Li, G., Xiong, W., Ma, J., and Shen, S. (2012) Dynamic activation of the key pathways: linking colitis to colorectal cancer in a mouse model. Carcinogenesis 33, 1375–1383

33. Park, J., and Hansen, C. G. (2021) Cellular feedback dynamics and multilevel regulation driven by the hippo pathway. Biochem Soc Trans 49, 1515–1527

34. Perez White, B. E., and Getsios, S. (2014) Eph receptor and ephrin function in breast, gut, and skin epithelia. Cell Adh Migr 8, 327–338

35. Grandi, A., Zini, I., Palese, S., Giorgio, C., Tognolini, M., Marchesani, F., Bruno, S., Flammini, L., Cantoni, A. M., Castelli, R., Lodola, A., Fusari, A., Barocelli, E., and Bertoni, S. (2019) Targeting the Eph/Ephrin System as Anti-Inflammatory Strategy in IBD. Front Pharmacol 10, 691

36. Darling, T. K., and Lamb, T. J. (2019) Emerging Roles for Eph Receptors and Ephrin Ligands in Immunity. Front Immunol 10, 1473

37. Arvanitis, D., and Davy, A. (2008) Eph/ephrin signaling: networks. Genes Dev 22, 416–429

38. Cai, J., Zhang, N., Zheng, Y., de Wilde, R. F., Maitra, A., and Pan, D. (2010) The Hippo signaling pathway restricts the oncogenic potential of an intestinal regeneration program. Genes Dev 24, 2383–2388

39. Schlegelmilch, K., Mohseni, M., Kirak, O., Pruszak, J., Rodriguez, J. R., Zhou, D., Kreger, B. T., Vasioukhin, V., Avruch, J., Brummelkamp, T. R., and Camargo, F. D. (2011) Yap1 acts downstream of alpha-catenin to control epidermal proliferation. Cell 144, 782–795

40. Karayiannakis, A. J., Syrigos, K. N., Efstathiou, J., Valizadeh, A., Noda, M., Playford, R. J., Kmiot, W., and Pignatelli, M. (1998) Expression of catenins and E-cadherin during epithelial restitution in inflammatory bowel disease. J Pathol 185, 413–418

41. Taniguchi, K., Wu, L. W., Grivennikov, S. I., de Jong, P. R., Lian, I., Yu, F. X., Wang, K., Ho, S. B., Boland, B. S., Chang, J. T., Sandborn, W. J., Hardiman, G., Raz, E., Maehara, Y., Yoshimura, A., Zucman-Rossi, J., Guan, K. L., and Karin, M. (2015) A gp130-Src-YAP module links inflammation to epithelial regeneration. Nature 519, 57–62

42. Yui, S., Azzolin, L., Maimets, M., Pedersen, M. T., Fordham, R. P., Hansen, S. L., Larsen, H. L., Guiu, J., Alves, M. R. P., Rundsten, C. F., Johansen, J. V., Li, Y., Madsen, C. D., Nakamura, T., Watanabe, M., Nielsen, O. H., Schweiger, P. J., Piccolo, S., and Jensen, K. B. (2018) YAP/TAZ-Dependent Reprogramming of Colonic Epithelium Links ECM Remodeling to Tissue Regeneration. Cell Stem Cell 22, 35–49 e37

43. Kuester, D., Guenther, T., Biesold, S., Hartmann, A., Bataille, F., Ruemmele, P., Peters, B., Meyer, F., Schubert, D., Bohr, U. R., Malfertheiner, P., Lippert, H., Silver, A. R., Roessner, A., and Schneider-Stock, R. (2010) Aberrant methylation of DAPK in long-standing ulcerative colitis and ulcerative colitis-associated carcinoma. Pathol Res Pract 206, 616–624

44. Yuan, W., Chen, J., Shu, Y., Liu, S., Wu, L., Ji, J., Liu, Z., Tang, Q., Zhou, Z., Cheng, Y., Jiang, B., and Shu, X. (2017) Correlation of DAPK1 methylation and the risk of gastrointestinal cancer: A systematic review and meta-analysis. PLoS One 12, e0184959

45. Lopes, F., Keita, A. V., Saxena, A., Reyes, J. L., Mancini, N. L., Al Rajabi, A., Wang, A., Baggio, C. H., Dicay, M., van Dalen, R., Ahn, Y., Carneiro, M. B. H., Peters, N. C., Rho, J. M., MacNaughton, W. K., Girardin, S. E., Jijon, H., Philpott, D. J., Soderholm, J. D., and McKay, D. M. (2018) ER-stress mobilization of death-associated protein kinase-1-dependent xenophagy counteracts mitochondria stress-induced epithelial barrier dysfunction. J Biol Chem 293, 3073–3087

46. Okamoto, M., Takayama, K., Shimizu, T., Ishida, K., Takahashi, O., and Furuya, T. (2009) Identification of death-associated protein kinases inhibitors using structure-based virtual screening. J Med Chem 52, 7323–7327

47. Al-Ghabkari, A., Deng, J. T., McDonald, P. C., Dedhar, S., Alshehri, M., Walsh, M. P., and MacDonald, J. A. (2016) A novel inhibitory effect of oxazol-5-one compounds on ROCKII signaling in human coronary artery vascular smooth muscle cells. Sci Rep 6, 32118

48. Davies, S. P., Reddy, H., Caivano, M., and Cohen, P. (2000) Specificity and mechanism of action of some commonly used protein kinase inhibitors. Biochem J 351, 95–105

49. Ishizaki, T., Uehata, M., Tamechika, I., Keel, J., Nonomura, K., Maekawa, M., and Narumiya, S. (2000) Pharmacological properties of Y-27632, a specific inhibitor of rho-associated kinases. Mol Pharmacol 57, 976–983

50. Ito, H., Yamashita, Y., Tanaka, T., Takaki, M., Le, M. N., Yoshida, L. M., and Morimoto, K. (2020) Cigarette smoke induces endoplasmic reticulum stress and suppresses efferocytosis through the activation of RhoA. Sci Rep 10, 12620

51. Li, K. X., Du, Q., Wang, H. P., and Sun, H. J. (2019) Death-associated protein kinase 3 deficiency alleviates vascular calcification via AMPK-mediated inhibition of endoplasmic reticulum stress. Eur J Pharmacol 852, 90–98

52. Lai, Y., Wu, B., Chen, L., and Zhao, H. (2004) A statistical method for identifying differential gene-gene co-expression patterns. Bioinformatics 20, 3146–3155

53. McKenzie, A. T., Katsyv, I., Song, W. M., Wang, M., and Zhang, B. (2016) DGCA: A comprehensive R package for Differential Gene Correlation Analysis. BMC Syst Biol 10, 106

